# Trends of SARS-CoV-2 antibody prevalence in selected regions across Ghana

**DOI:** 10.1101/2021.04.25.21256067

**Authors:** Peter Kojo Quashie, Joe Kimanthi Mutungi, Francis Dzabeng, Daniel Oduro-Mensah, Precious C. Opurum, Kesego Tapela, Aniefiok John Udoakang, WACCBIP COVID-19 Team, Ivy Asante, Lily Paemka, Frederick Kumi-Ansah, Osbourne Quaye, Emmanuella Amoako, Ralph Armah, Charlyne Kilba, Nana Afia Boateng, Michael Ofori, George B. Kyei, Yaw Bediako, Nicaise Ndam, James Abugri, Patrick Ansah, William K. Ampofo, Francisca Mutapi, Gordon A. Awandare

## Abstract

To estimate the level of community exposure to SARS-CoV-2 in Ghana, we conducted phased seroprevalence studies of 2729 participants in selected locations across Ghana. Phase I screening (August 2020) covered a total of 1305 individuals screened at major markets/lorry stations, major shopping malls, hospitals and research institutions involved in COVID-19 work. The screening was performed using a strip-in-cassette lateral flow type Rapid Diagnostic Test (RDT) kit that simultaneously and separately detected IgM and IgG antibodies against SARS-CoV-2 nucleocapsid protein. In Phase I, 252/1305 (19%) tested positive for IgM or IgG or both. Exposure rate was significantly higher among individuals tested at markets/lorry stations (26.9%) compared to those at Shopping Malls (9.4%). The 41–60-years age group had the highest exposure rate (27.2%). People with only a basic level or no formal education had a higher exposure rate (26.2%) than those with tertiary level education (13.1%); and higher in informally employed workers (24.0%) than those in the formal sector (15.0%). Phases II and III screening activities in October and December 2020, respectively, showed no evidence of increased seroprevalence, indicating either a reduced transmission rate or loss of antibody expression in a subset of the participants. The Upper East region has the lowest exposure rate, with only 4 of 200 participants (2%) seropositivity. Phase IV screening in February 2021 showed that exposure rates in the upper income earners (26.2%) had almost doubled since August 2020, reflective of Ghana’s second wave of symptomatic COVID-19 cases, which began in December 2020. The Phase IV results suggest that seroprevalence levels have become so high that the initial socioeconomic stratification of exposure has been lost. Overall, the data indicates a much higher COVID-19 seroprevalence in the Greater Accra Region than was officially acknowledged, likely implying a considerably lower case fatality rate than the current national figure of 0.84%. Additionally, the high exposure levels seen in the communities suggest that COVID-19 in Ghana still predominantly presents with none-to-mild symptoms. Our results lay the foundation for more extensive SARS-CoV-2 surveillance in Ghana and the West African sub-region, including deploying rapid antigen test kits in concert to determine the actual infection burden since antibody development lags infection.

## Background

The severe acute respiratory syndrome coronavirus 2 (SARS-CoV-2) was first reported in Wuhan, China, in late 2019 [1]. By April 20^th^ 2021 there were 141,058,320 COVID-19 SARS-CoV-2 reported infections with 3,015,314 associated deaths (case fatality ratio (CFR): of 2.1%) globally. Of these, 4,437,846 COVID-19 cases and 118,133 deaths (CFR: 2.7%), representing 3.07% and 3.85% of all reported global cases and deaths, were from the 55 African Union Member States [2]. Ghana, from the first two reported (imported) cases on March 12^th^ 2020, by April 16^th^ 2021 reported a cumulative total of 91,709 confirmed cases with 771 associated deaths (CFR of 0.84%) by this date [3].

The current gold standard method for diagnosis of SARS-CoV-2 infection is by real-time reverse transcription polymerase chain reaction (RT-PCR), which detects viral nucleic acid sequences, and thus the virus, present in the upper respiratory tract (nasopharyngeal or oropharyngeal) swab samples [4-6]. Due to limited resources, tests are prioritised on symptomatic, severe, and/or suspected cases, and occasionally on contacts of confirmed cases. Common symptoms of SARS-CoV-2 infection include fever, dry cough, tiredness and other variable ones with onset between 5 to 14 days after infection [7]. Approximately 80% of infected persons show mild or no symptom [8], however, posing a danger of unmitigated transmission and potential rapid rise in disease onset, severity and death [9]. Additionally, RT-PCR sensitivity may be affected by viral load, virus replication rate, RNA isolation method, and the source or timing of swab collection relative to disease stage [10]. This could lead to false negativity of about 20% [11], indicating that actual infections may be higher than reported per test.

Rapid immunodiagnostic tests (RDTs) can be used to detect either SARS-CoV-2-expressed proteins (antigens) in respiratory tract samples (e.g., sputum, throat swab) or human anti-SARS-CoV-2 specific antibodies in blood or serum within minutes. Rapid antigen tests have utility for rapid detection of transmissible infections but have lower sensitivity than PCR-based methods [12] and have become common for routine SARS-CoV-2 screening at ports of entry. Some RDTs also detect the presence of antibodies against SARS-CoV-2 in bodily fluids. These are useful for performing population level surveillance of viral exposure. Unlike antigen RDT’s which pick up active and often transmissible infection, antibody RDTs tend to pick up the evidence of previous or recent infection and cannot be used to detect active infection [12]. There are more than 280 Conformité Européenne-*in vitro* diagnostics (CE-IVD)-marked COVID-19 antibody detection RDT kits listed with the Foundation for Innovative Diagnostics (FIND) [13]. Currently, lateral flow immunoassays (LFIAs), chemiluminescence immunoassays (CLIAs), or enzyme-linked immunosorbent assays (ELISAs) are commonly used for detection of SARS-CoV-2 IgM and IgG antibodies [14, 15]. Similar to other infections, SARS-CoV-2 elicits an immune response [16], and the presence of the virus-specific antibodies in blood indicates previous or current infection regardless of the presence or absence of symptoms [17, 18]. Generally, IgM or IgG are produced early and later in infection, respectively [19]. It is currently unknown how long SARS-CoV-2 IgM and IgG antibodies persist, however, seroconversion of IgM appears to peak simultaneously with IgG within 2 to 3 weeks after symptoms onset [10, 17, 20-24]

In addition to retrospectively evaluating infection dynamics and the population disease burden, serology tests are useful for vaccine trials, therapeutic antibodies results analyses, and tests for individual and herd immunity. Antibody presence can also help identify COVID-19-recovered individuals and potential donors of convalescent plasma for immunotherapy of critically sick COVID-19 patients [25]. Some seroprevalence studies have used cross-sectional snapshots to evaluate community level exposure of SARS-CoV-2 [26] but to our knowledge, no single study has attempted to track the spatial-temporal dynamics of SARS-CoV-2 exposure in Africa. Ghana reported its first two (imported) cases on March 12^th^, 2020 [3]. By April 4^th^, positive cases who had neither travel history nor known contact with confirmed cases, were detected, implying local transmission. To estimate COVID-19 community spread in Ghana, over a 7-month period, we randomly screened for IgM and IgG antibodies against SARS-CoV-2 in people at various public places in Accra (National capital, where >50% of reported cases occur), Kasoa (a densely-populated town in the Central Region, and a COVID-19 transmission hotspot [27] and which shares a border with Accra), Cape Coast (Central Regional capital), Akropong (a small town in the Eastern Region, a 15-minute drive away from Accra), Navrongo (a small town in the Upper East Region, which hosts a public university and a government Health Research Centre) and Bolgatanga (the Upper East Regional capital) (Figure 1). A questionnaire administered during the study collected demographic data as well and evaluated the COVID-19 knowledge, attitudes and perceptions (KAP) of study participants.

**Figure 1:**
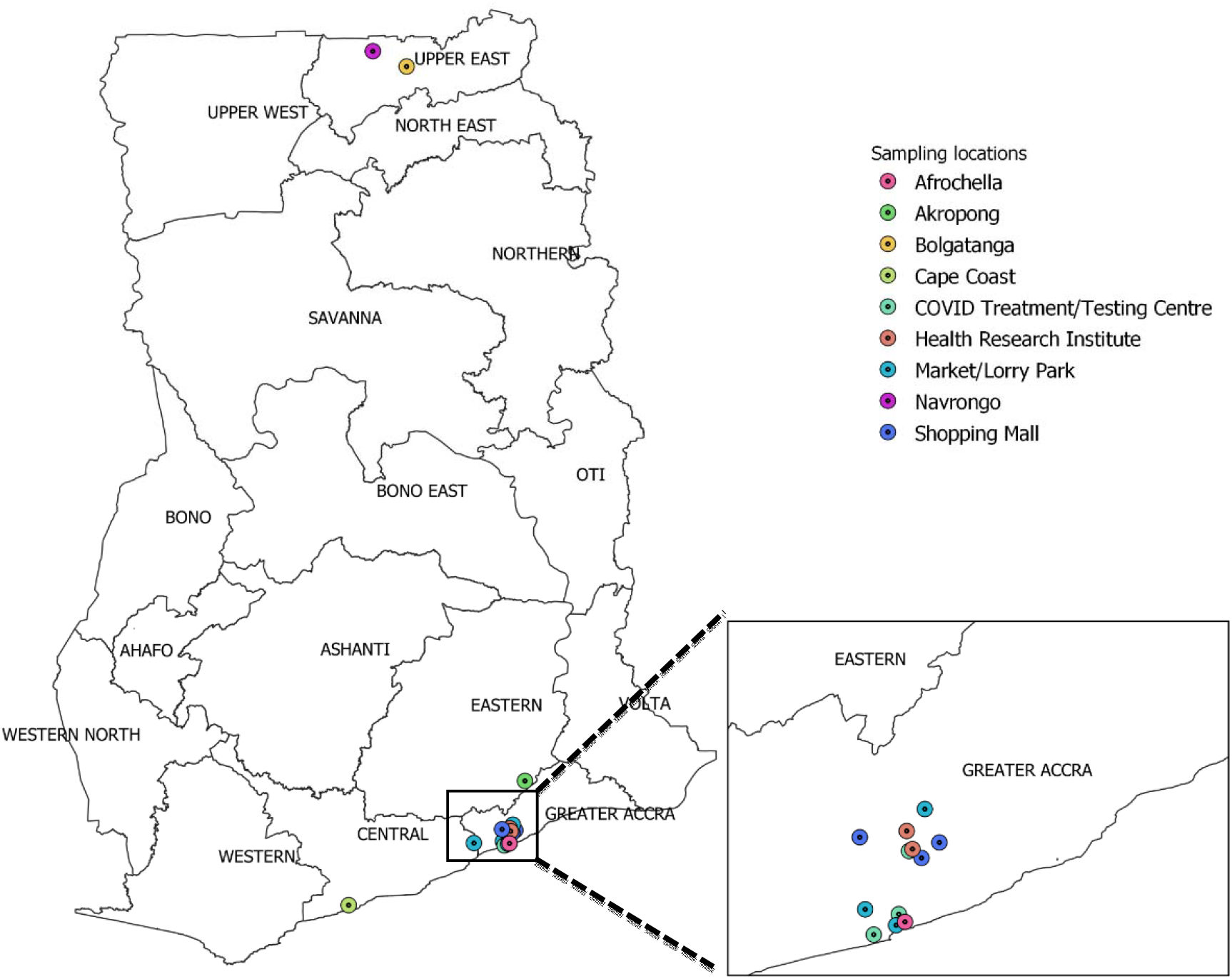
Map of Ghana showing study sites. Figure was generated using QGIS (QGIS Development Team, 2009. QGIS Geographic Information System. Open Source Geospatial Foundation. URL http://qgis.org)

## Methods

### Study design

This study was a multi-site repeated observational cross-sectional study carried out over a period of 7 months from July 27, 2020 to February 26, 2021. Phase I was performed between 27 July and September 14, 2020 (designated August 2020), followed by additional phases in October 2020, December 2020 and February 2021 to identify changes from the initial rates observed at public places (Figure S1). For ease of reference in the text, and for site anonymity, the sites were assigned codes based on site type and risk factors: markets and lorry stations (ML), malls (M), research centres (R), hospitals (H) and generalised community screening (C). Generalised community screening included attendees at a concert since that reflected individuals who would be otherwise dispersed through the community. A research centre involved in mass COVID-19 testing and a COVID-19 treatment centre were given the respective codes, RC and HC. Phase I participants were invited to volunteer for the study at two shopping malls, three major markets/lorry stations (ML1-3), two research institutes involved in COVID-19 work (R1) and COVID-19 testing (RC), and three major hospitals, one of which was a COVID-19 treatment centre (H1, H2, and HC). Informed consent was obtained from all study participants. Exposure to COVID-19 was detected using a strip-in-cassette lateral flow rapid diagnostic test kit which simultaneously detects IgM and IgG antibodies against SARS-CoV-2 antigens. Phase II screened participants at one market (ML4), one research centre (R1) and two hospitals (H1, H4) while Phase III screened participants at ML1 and across two towns in the Upper East Region (C1). During Phase IV, the exposure levels of upper-income earners were evaluated by screening at 2 malls (M1 and M3) and a repeat screening at H2. In addition, the exposure level in a small town (C3) in the Eastern Region, near Accra, was estimated. Screening at hospitals and research facilities included only staff members and their close contacts; patients at the hospitals were excluded from this study. All tests were performed on-site and participants were subsequently informed of their exposure status and counselled to adhere to COVID-19 mitigation protocols. When IgM was detected, participants were referred for a COVID-19 PCR test.

### Testing kit

The ‘UNSCIENCE COVID-19 IgG/IgM antibody Rapid Test Kit’ (Catalogue# UNCOV-40, Lot Number 20200326) was registered with the Ghana FDA, and the kit validation report was submitted to the Ghana Food and Drug Authority (FDA). For the validation exercise, sera from RT-PCR confirmed COVID-19 cases were used to evaluate several kits’ performance since there were no established SARS-CoV-2 antibody detection standards for use at the commencement of this study. This kit had a manufacturer-declared IgG sensitivity and specificity of over 98% (https://covid-19-diagnostics.jrc.ec.europa.eu/devices/detail/634). Using the Ghana Food and Drugs Authority (FDA) validation protocol^36^, the UNSCIENCE kit demonstrated a sensitivity of 66% when tested using 100 Ghanaian convalescent COVID-19 patient sera (2-4 weeks after a PCR-positive result). We checked the existence of pre-existing cross-reactive antibodies using sera from 100 PCR-verified COVID-19 negative samples and obtained a specificity of 94%. In the validation exercises, when a test result was not obvious, at least 3 researchers validated the reading. In the rare case of an invalid test (no control line, or wrong location of bands), the test was repeated. A representative set of randomly chosen positive and negative test results are shown in Figure S2. The kit was also adjudged to have a concordance of 72% with the WHO-recommended Wantai ELISA kit (https://www.fda.gov/media/140929/download).

### Data management and analysis

A short questionnaire was administered to capture participant demographic data, knowledge of COVID-19 and COVID-19 testing history. The data, including the antibody test results were entered and managed using <u>R</u>esearch <u>E</u>lectronic <u>D</u>ata <u>Cap</u>ture suite (REDCap) [28]. The data were cleaned by checking for completeness, duplication and consistency. Cleaned data were analyzed with Stata 16 (StataCorp, College Station, Texas, USA) and R/Rstudio [29, 30]. GraphPad Prism version 8.0.0 [31] was used for some additional analysis and generation of figures. Descriptive analyses were performed and univariate and multivariate logistic regression models were used to assess the association between seroprevalence and risk factors. Multivariable logistic models for seropositivity were obtained by using a backward stepwise procedure. Demographic variables that were associated with seropositivity at the P<0.25 level were included. The overall goodness of fit was assessed using the Wald statistic. Unadjusted and adjusted odds ratios with 95% confidence intervals were computed and presented as parallel dot plots with error bars. Statistical significance was inferred for *p*-values below 0.05.

## Results

### Study sites and participant characteristics

A total of 2729 participants were screened for this study (Figure S1), including 1305 in Phase I (10 sites: August, 2020), 395 in Phase II (4 sites: October, 2020), 393 in Phase III (2 sites: December, 2020) and 636 in Phase IV (4 sites: February 2021). Altogether, four markets/lorry stations (ML-1 was visited twice), three malls (M1, twice), two research institutes (R, twice), two sets of small towns locations (C1 and C3) and four hospitals (H1 and H2, twice) were screened across all Phases. During Phase III, 200 individuals were also screened from Navrongo and Bolgatanga in the Upper East Region (C1), which had the lowest reported COVID-19 cases in Ghana at the time the study began. Attendees (81) at a free Afrochella concert in Accra were also screened during Phase III (C2), potentially representing a general cross-section of the GAR population.

During Phase I, 946 individuals were sampled in public spaces, including 616 at markets/lorry stations and 330 at malls, while 359 were sampled in healthcare (254) and research (105) facilities (Table 1). There was a slightly higher number of female participants than males. The modal age range was 21–40 years, and over 40% of the participants had a tertiary education. Tertiary education was defined as having a post-secondary qualification such as a diploma, degree or above. Most respondents worked in the informal sector, with low or mid-level socioeconomic status. Over 90% of participants had good knowledge of COVID-19 symptoms, transmission routes and preventative measures (Figure S3). Such knowledge did not, however, correlate with participants’ seropositivity status or mask-wearing prior to recruitment. Only 7% of participants had previously received a COVID-19 PCR test (Figure S4). Similar participant characteristics were observed in Phases II, III and IV with smaller numbers of participants.

**Table 1:** Socio-demographic characteristics of participants (N=2729)

| Characteristics | Phase 1 (n=1305) |  | Phase 2 (n=395) |  | Phase 3 (n=393) |  | Phase 4 (n=636) |  |
| --- | --- | --- | --- | --- | --- | --- | --- | --- |
|  | number | Percentage | number | Percentage | number | Percentage | number | Percentage |
| **Exposure risk groups** |  |  |  |  |  |  |  |  |
| Shopping malls (M) | 330 | 25.3 |  |  |  |  | 356 | 56.0 |
| Markets/Lorry stations (ML) | 616 | 47.2 | 152 | 38.5 | 115 | 36.5 |  |  |
| COVID testing/treatment centres (RC/HC) | 105 | 8.0 |  |  |  |  |  |  |
| Other Health Research centres/Hospital R/H | 254 | 19.5 | 243 | 61.5 |  |  | 81 | 12.7 |
| Navrongo/Bolgatanga (C1) |  |  |  |  | 200 | 63.5 |  |  |
| Afrochella Concert (C2) |  |  |  |  | 78 | 19.8 |  |  |
| Akropong (C3) |  |  |  |  |  |  | 199 | 31.3 |
| **Gender** |  |  |  |  |  |  |  |  |
| Male | 591 | 45.3 | 124 | 31.4 | 177 | 45.0 | 295 | 46.4 |
| Female | 714 | 54.7 | 271 | 68.6 | 216 | 55.0 | 341 | 53.6 |
| **Age group (in years)** |  |  |  |  |  |  |  |  |
| <21 | 63 | 4.8 | 13 | 3.3 | 63 | 16.0 | 44 | 6.9 |
| 21-40 | 769 | 58.9 | 216 | 54.7 | 251 | 63.9 | 362 | 56.9 |
| 41-60 | 365 | 28.0 | 119 | 30.1 | 57 | 14.5 | 167 | 26.3 |
| 60+ | 89 | 6.8 | 13 | 3.3 | 7 | 1.8 | 62 | 9.7 |
|  | number | Percentage | number | Percentage | number | Percentage | number | Percentage |
| Missing | 19 | 1.5 | 34 | 8.6 | 15 | 3.8 | 1 | 0.2 |
| **Educational level** |  |  |  |  |  |  |  |  |
| None/Basic | 402 | 30.8 | 128 | 32.4 | 117 | 29.8 | 202 | 31.8 |
| Secondary | 245 | 18.8 | 54 | 13.7 | 148 | 37.7 | 139 | 21.9 |
| Tertiary | 616 | 47.2 | 133 | 33.7 | 106 | 27.0 | 279 | 43.9 |
| Missing | 42 | 3.2 | 80 | 20.3 | 22 | 5.6 | 16 | 2.5 |
| **Employment type** |  |  |  |  |  |  |  |  |
| Formal | 520 | 39.8 | 171 | 43.3 | 72 | 18.3 | 257 | 40.4 |
| Informal | 655 | 50.2 | 175 | 44.3 | 190 | 48.3 | 261 | 41.0 |
| Student | 59 | 4.5 | 21 | 5.3 | 114 | 29.0 | 48 | 7.5 |
| Unemployed | 44 | 3.4 | 19 | 4.8 | 11 | 2.8 | 70 | 11.0 |
| Missing | 27 | 2.1 | 9 | 2.3 | 6 | 1.5 | 0 | 0.0 |
| **Socio-economic status** |  |  |  |  |  |  |  |  |
| Lowest | 524 | 40.2 | 170 | 43.0 | 158 | 40.2 | 249 | 39.2 |
| Middle | 259 | 19.8 | 122 | 30.9 | 78 | 19.8 | 124 | 19.5 |
| Higher | 522 | 40.0 | 103 | 26.1 | 157 | 39.9 | 263 | 41.4 |

### Anti-SARS-CoV-2 seropositivity in Phase I

In Phase I, SARS-CoV-2 IgG, IgM or both antibodies were detected in 19% of all participants (Figure 2A), with the highest rate amongst participants sampled in markets/lorry stations (27%). Among health workers, those at COVID-19 treatment/testing sites had higher exposure rates compared to their colleagues who were not directly handling COVID-19 patients or samples. There was no significant difference in seropositivity across genders (Figure 2B). When stratified by age categories, the highest level of seroprevalence (27.1%) was observed in the 41–60 years age group (Figure 2C). Participants with higher educational backgrounds (Figure 2D), those employed in the formal sector (Figure 2E) and those with higher economic standing (Figure 2F) had lower exposure levels than participants with lower educational background, informal sector workers and poor economic background. Only 20.9% of seropositive participants reported having had COVID-19-like symptoms (Figure 3).

**Figure 2:**
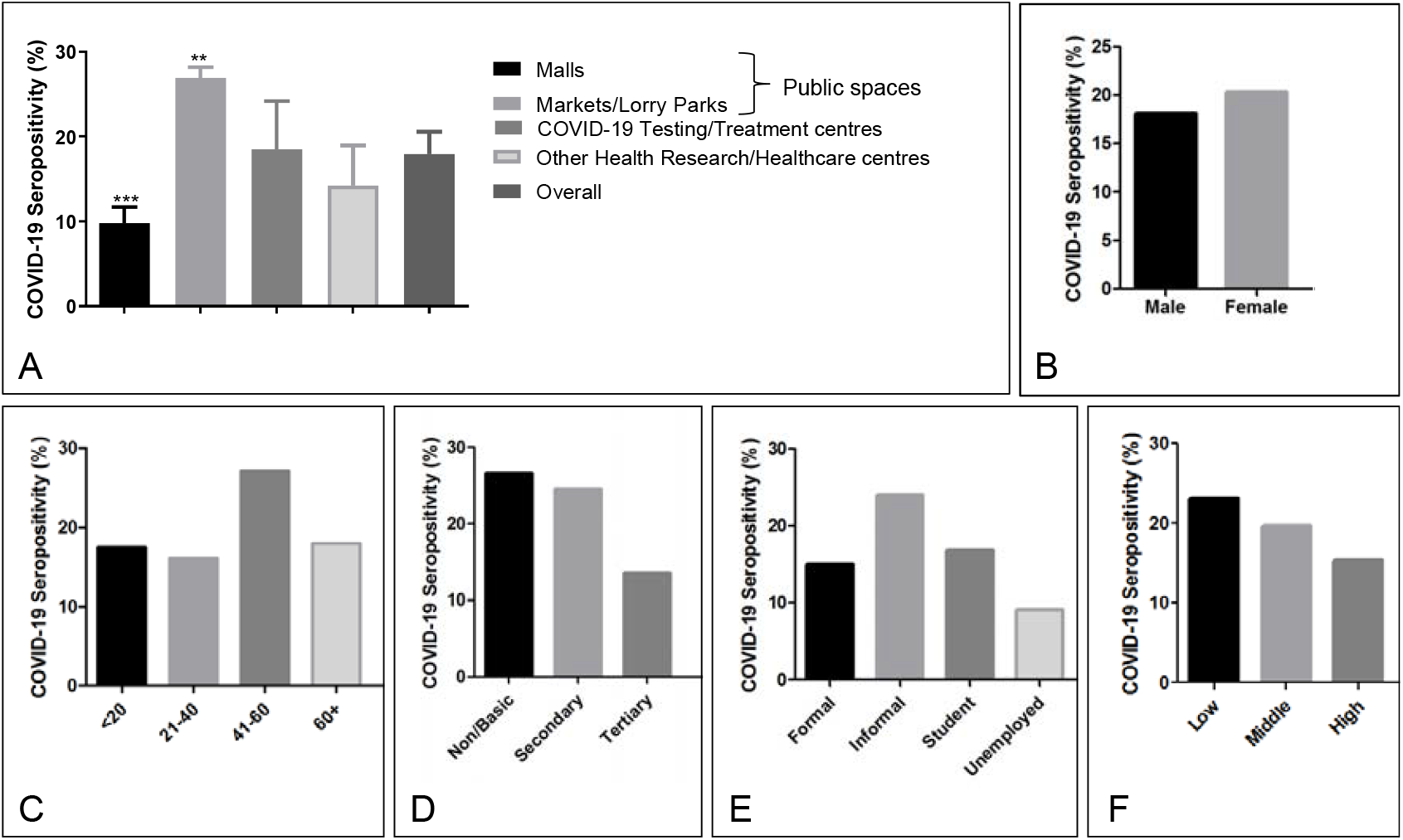
SARS-CoV-2 seropositivity reported by (A) Sampling sites, (B) gender, (C) Age, (D) Highest education level, (E) Employment status and (F) Socioeconomic status. Error bars reflect the standard error of measurement. Where relevant, p-values are indicated as follows p<0.05; *,p<0.01;**, p<0.001; ***. Error bars, where relevant, represent standard deviation across sites.

**Figure 3.**
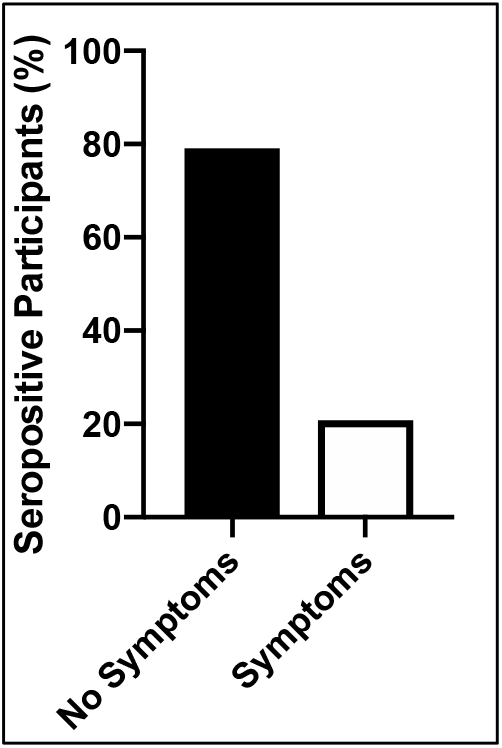
Presence of two or more self-reported COVID-19 symptoms in seropositive individuals in the month preceding the study.

Logistic regression analysis was performed on data from Phase I to identify factors that correlated with increased risk of SARS-CoV-2 exposure (Figure 4). Univariate modelling showed that socio-demographic factors were significantly associated with increased or decreased exposure. These were: being sampled at either markets and lorry stations (Odds ratio, OR:3.6, 95% confidence interval, CI: 2.4-5.4) or a COVID-19 treatment/testing centre (OR: 2.4, 95% CI:1.3-4.4,), being employed in the informal sector (OR: 1.8, 95% CI:1.3-2.4) having a high education (OR: 0.4, 95% CI:0.3-0.6) or having a high income (OR:0.6, 95% CI:0.5-0.9). In a multivariate model, sampling location, participant educational level and income/socioeconomic status remained significantly associated with COVID-19 exposure status; participants were significantly more likely to have COVID-19 antibodies if they were sampled in markets/lorry stations (adjusted odds ratio, aOR: 2.5, 95% CI: 1.5-4.2), and COVID-19 testing/treatment centre (aOR:3.6, 95%CI:1.7-7.5). Participants who had basic or no formal education (aOR: 0.8, 95% CI: 0.5-1.3) had a higher risk of COVID-19 seropositivity. Unemployed individuals (aOR: 0.7, 95% CI: 0.2-2.1) (Table 2, Figure 4), those with high educational level (aOR: 0.8, 95%CI: 0.5-1.3) and high socioeconomic status (aOR: 0.8, 95%CI: 0.6-1.2) had reduced risk of COVID-19 seropositivity, but these associations were not statistically significant (Table 2, Figure 4).

**Table 2:** Summary of participant seropositivity status (N=2729)

|  | August 2020 (n=1305) |  | Oct 2020 (n=395) |  | Dec - 2020 (n=393) |  | Jan- Feb 2021 (n=636) |  |
| --- | --- | --- | --- | --- | --- | --- | --- | --- |
|  | Number | Antibody Positivity | Number | Antibody Positivity | Number | Antibody Positivity | Number | Antibody Positivity |
|  | Sampled | % | Sampled | % | Sampled | % | Sampled | % |
| **Sampling location (Code)** |  |  |  |  |  |  |  |  |
| Shopping malls (M) | 330 | 9.4 |  |  |  |  | 356 | 25.3 |
| Markets/Lorry stations (ML) | 616 | 26.9 | 152 | 19.7 | 115 | 23.5 |  |  |
| COVID testing/treatment Centres (RC/HC) | 105 | 20.0 |  |  |  |  |  |  |
| Health Research Centres/Hospital (R/H) | 254 | 13.4 | 243 | 14.0 |  |  | 81 | 22.2 |
| Navrongo/Bolgatanga (C1) |  |  |  |  | 200 | 2.0 |  |  |
| Afrochella Concert (C2) |  |  |  |  | 78.0 | 15.4 |  |  |
| Akropong (C3) |  |  |  |  |  |  | 199 | 18.6 |
| **Gender** |  |  |  |  |  |  |  |  |
| Male | 591 | 18.1 | 124 | 14.5 | 131 | 1.7 | 10.7 | 22.7 |
| Female | 714 | 20.3 | 271 | 17.0 | 184 | 2.8 | 11.1 | 22.9 |
| **Age group (in years)** |  |  |  |  |  |  |  |  |
| 4-21 | 63 | 17.5 | 13 | 0.0 | 63 | 11.1 | 44 | 9.1 |
| 21-40 | 769 | 16.1 | 216 | 11.1 | 251 | 11.6 | 362 | 24.9 |
| 41-60 | 365 | 27.1 | 119 | 21.8 | 57 | 10.5 | 167 | 22.8 |
| 60+ | 89 | 18.0 | 13 | 38.5 | 7.0 | 14.3 | 62 | 21.0 |
| Missing | 19 | 10.5 | 34 | 26.5 | 15 | 0.0 | 1 | 0.0 |
|  | Number | Antibody Positivity | Number | Antibody Positivity | Number | Antibody Positivity | Number | Antibody Positivity |
|  | Sampled | % | Sampled | % | Sampled | % | Sampled | % |
| **Educational level** |  |  |  |  |  |  |  |  |
| None/Basic | 402 | 26.6 | 128 | 22.7 | 117 | 12.0 | 202 | 17.8 |
| Secondary | 245 | 24.5 | 54 | 14.8 | 148 | 11.5 | 139 | 25.9 |
| Tertiary | 616 | 13.5 | 133 | 15.0 | 106 | 10.4 | 279 | 24.7 |
| Missing | 42 | 4.8 | 80 | 8.8 | 22 | 4.5 | 16 | 25.0 |
| **Employment type** |  |  |  |  |  |  |  |  |
| Formal | 520 | 15.0 | 171 | 11.7 | 72 | 6.9 | 257 | 23.7 |
| Informal | 655 | 24.0 | 175 | 20.6 | 190 | 13.7 | 261 | 24.9 |
| Student | 59 | 16.9 | 21 | 4.8 | 114 | 9.6 | 48 | 12.5 |
| Unemployed | 44 | 9.1 | 19 | 21.1 | 11 | 0 | 70 | 18.6 |
| Missing | 27 | 11.1 | 9 | 33.3 | 6 | 16.7 |  |  |
| **Socio-economic status** |  |  |  |  |  |  |  |  |
| Low | 524 | 23.1 | 170 | 17.1 | 158 | 15.8 | 249 | 18.1 |
| Middle | 259 | 19.7 | 122 | 15.6 | 78 | 10.3 | 124 | 25.0 |
| High | 522 | 15.3 | 103 | 15.5 | 157 | 6.4 | 263 | 26.2 |
|  | IgG | 13.0 | IgG | 15.2 | IgG | 9.7 | IgG | 22.8 |
|  | IgM | 1.6 | IgM | 1.3 | IgM | 2.3 | IgM | 2.2 |

**Figure 4:**
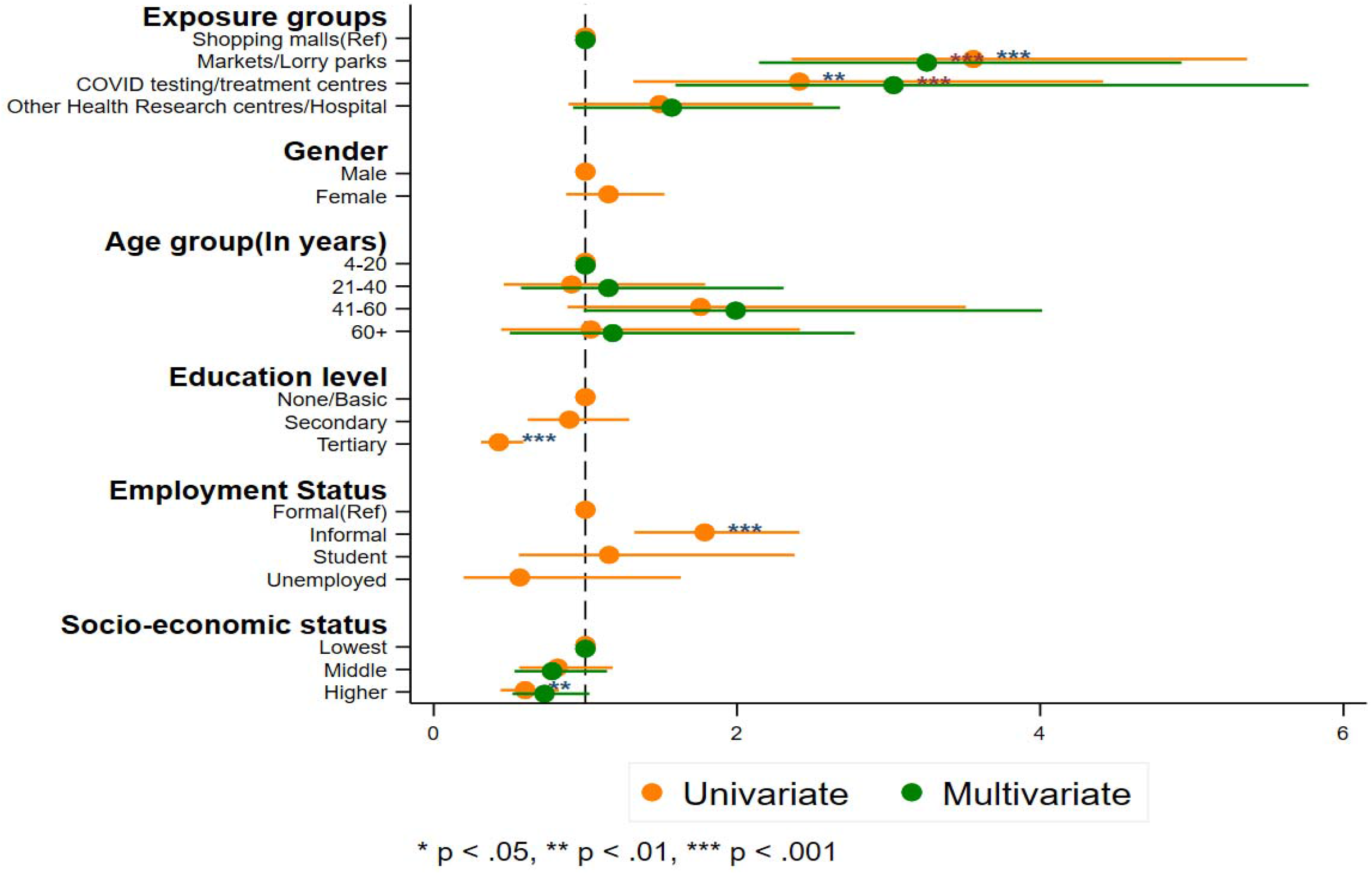
Modelling of COVID-19 exposure risk across Phase I study participant.

### Phases II and III: Targeted follow-up seroprevalence surveys

As follow-ups to Phase I, the trend of population seroprevalence was again investigated in Phase II and III. Two months after the initial public places screening (October, 2020), 144 individuals were screened at a lorry station in Accra and 212 participants at two hospitals in Accra and Cape Coast. Overall, seroprevalence at the lorry station was 19.7%, and 13% at the two hospitals. The H4 facility staff had higher (18.5%) seroprevalence level than those at H1, previously sampled in Phase I. A Phase III screening exercise was conducted at M1, which was originally sampled in Phase I, and that showed an estimated seroprevalence of 23.5%. Additionally, 200 individuals were screened in the Upper East Region (C1), an area with low population density and very few reported COVID-19 cases at the time. Here, 4 out of 200 individuals (2%) tested positive for SARS-CoV-2 antibodies (Table 1). Individuals screened at the Afrochella concert (C2) showed an estimated seroprevalence of 16%, which was similar to the Phase I data, with the HC site excluded.

### Phase IV: Impact of the second wave

Beginning in late December 2020, large numbers of symptomatic COVID-19 cases started to be detected at hospitals and treatment centres in Accra and other major cities. The patients were mostly of high socio-economic standing [32]. We therefore performed repeat screening at M1 and H2 (both sampled in Phase I) and sampled again at M3 and a small town in the Eastern Region (C3). The average seroprevalence at the two malls (M1 and M3) was at 27%, whilst H1 and C3 recorded 25% and 17% respectively.

## Discussion

This study was necessitated by a dearth of epidemiological data on COVID-19 prevalence in Ghana. In the first few months of the pandemic when prevalence was low, Ghana ranked high among African countries, and even globally, for administering high numbers of tests per million people [33]. To meet the high demand for testing, Ghana’s main testing centre, Noguchi Memorial Institute for Medical Research, employed “sample pooling” methods [34-37]. However, since then, Ghana has declined significantly to number 22 in tests per million of population in Africa [38]. Data from the Ghana Health Service’s COVID-19 archives [3] indicates that testing has significantly reduced after peaking in June, correlating with a drop in daily reported cases. Among other factors, the reduced testing could be due to the fact that at the current positivity rate of 8.3% of tested cases, sample pooling is no longer a viable cost-cutting and test-rate enhancing measure. The seroprevalence rate average of 19.3% obtained from our public screening exercises is probably a better reflection of SARS-CoV-2 infections in Ghana, especially in the large and densely populated urban areas. Additionally, currently, most RT-PCR tests in the country are administered to travellers, representing a higher economic tier of society. The relatively low (9.3%) seroprevalence initially observed in malls, assumed to be frequented by the higher tiers of society, may correlate well with the official 10% RT-PCR test positivity rate reported in September 2020 [3].

Participants across all sites demonstrated good knowledge of COVID-19 risks, symptoms and preventive measures. This did not however translate into observation of protocols in the markets and lorry stations, where, by visual estimation, 10–50% of the study participants arrived mask-less and had to be requested to wear a mask donated by the study. This attitude corresponds with two surveys on mask-wearing, carried out by the Ghana Health Service which showed public mask wearing of ∼40% and 10% in July and September, respectively [39, 40]. Our previous genomic study showed evidence of undetected community spread likely caused by asymptomatic individuals [27]. Of note, nearly 80% of people who were seropositive did not report significant COVID-19 symptoms (Figure 3), confirming that SARS-CoV-2 infections in Ghana are predominantly asymptomatic, consistent with reported global trends [8]. With the 19.3% seroprevalence in the Greater Accra Region (GAR), we inferred that nearly 1 million out of the estimated 5 million GAR residents may have already been exposed to SARS-CoV-2. This staggering number suggests that the actual fatality rate of COVID-19 in Ghana may be much lower than the reported CFR of 0.7%, since that would have translated to approximately 7000 deaths in a large metropolis like Accra, which would have been rather very obvious. Additionally, there was no evidence of a stressed or panicked healthcare system nor visible or anecdotal evidence of excess deaths during the Phase I hospital screening.

Given the higher-than-expected seroprevalence observed in the Greater Accra Region, follow-up surveys were conducted at some of the Phase I locations as well as other parts of the country to confirm the findings and obtain vital information about infection dynamics over time. Repeated screening of markets and lorry stations in October and December yielded seroprevalence rates of 19.7% and 23.5%, respectively. The seeming plateau in seroprevalence in lower socioeconomic status individuals may be indicative of, either a peaking of infections -coincident with the onset of Phase I seroprevalence [3], or loss of antibody expression in some section of the population over time. Phase II surveys at the health and research facilities which were not directly handling patients or testing samples, in October, also showed a similar overall seroprevalence (13%), as observed in August. Participants in Navrongo and Bolgatanga in the Upper East region (C1), with some of the lowest reported cases, showed very low (2%) seroprevalence, confirming the reliability of this study. Consistent with global reports, participants in the 40-60 and 60+ year age groups exhibited the highest seroprevalence levels across all 3 phases (Figure 4, S4, S5). The results of Phase IV reflected the high levels of new COVID-19 cases and hospitalisations in Ghana, mainly in the middle socioeconomic class [32]. As such, it was not surprising that the socioeconomic divide in prevalence observed in Phase I had mostly disappeared and even appeared reversed by Phase IV.

Our observed seropositivity rates are in line with previous reports from other African countries[41, 42]. A study in Kenya estimated 20% SARS-CoV-2 seropositivity in adults (∼1.6 million people) at a time when the total reported infections were 2093 (with approximately 90% asymptomatic cases) and 71 deaths of all ages [43]. The initial trend relating income disparity and COVID-19 seropositivity is not surprising. Even in countries where lower seroprevalence was reported, such as China (1.63%), lower income status correlated with the highest seroprevalence (5.62%) [44], and this trend was initially reflected in this current study. The shift to high prevalence even in high socioeconomic brackets is likely due to poor adherence to COVID-19 protocols during the period of this study, likely due to election activities [45] and the 2020 Christmas/2021 New Year holiday festivities.

### Strengths and Limitations of this study

By surveying participants at different sites and times, representing categories of different perceived risk factors, this study obtained credible estimates for population-level prevalence across these sites and how that changed over the sampling period. This will allow future screening at these sites to determine the seroprevalence trends. However, most seroprevalence studies only reflect past disease burden. Using Markets and Lorry Stations enabled sampling of a broad cross-section of the Ghanaian populace.

Phase I of the study was conducted in the region with the greatest burden of reported infections and it was expected that a country-wide survey would yield less seroprevalence. Site H3, situated in the town of Cape Coast, a tourist hub and Central regional capital, exhibited a very high prevalence at 18.5% during Phase II, but this was not surprising given that M4, located in Kasoa, also in the Central Region, exhibited an exposure rate of 28% during Phase I. The low seroprevalence observed at C1 (2%) during Phase III hinted that community size/density may play a role in COVID-19 transmission. Given the geographical remoteness of C1 to the major hotspots of Accra and Kumasi, another small community (C3) in the country’s Southern belt with higher population density was screened, yielding an observed prevalence rate of 17%, and showing that SARS-CoV-2 exposure is not just a metropolitan burden, but one that needs to be tracked across the country. This, however, does not rule out that towns with lower population densities and who are far from metropolitan areas may exhibit lower seroprevalence levels.

During validation, this kit showed a sensitivity of 66%, when compared to PCR positivity. This is despite the manufacturer reporting sensitivity and specificity values above 98%. The apparent lower than expected sensitivity observed in local validation could at least partly be due to weak or delayed antibody responses in some of the infected persons. The import of this is that the seroprevalence levels reported in this study are likely underestimates of true disease prevalence.

One oft-repeated concern with SARS-CoV-2 seroprevalence studies in Africa is cross-reactivity due to pre-existing antibodies to other viruses and vaccines [46]. Some studies have reported extensive cross-reactivity against SARS-CoV-2 in Africa [47]). However, most of these studies had limitations, and as such their conclusions are unreliable. These flaws include extremely small study sizes (below 500 and even sometimes below 100) [48], tend to be based at single institutions and/or cities [47] and use samples collected at widely divergent time periods for their ‘Western’ and ‘African’ pre-COVID-19 samples [48]. The UNSCIENCE COVID-19 IgG/IgM antibody Rapid Test Kit used in our study exhibited 94% specificity during validation (with plasma from 100 COVID-19 PCR-negative individuals). Antibody RDTs are contraindicated in cases of active fever, based on manufacturer’s information leaflets, unpublished analyses and other studies [47]. We confirmed that none of the study respondents had temperature above 38, thereby reducing the likelihood of fever affecting the results.

Antibody cross-reactivity with other pathogens is often manifested in IgM detection [49, 50]. During validation, and in the field, detection of IgM was uncommon, and when detected, IgM was often accompanied by IgG. This reduced the likelihood that those IgM detections were as a result of cross-reactivity. That said, a cross-reactivity rate of 6% with IgM was detected during validation. However, the test kit performed even better in the field; Navrongo and Bolgatanga in the Upper-East region of Ghana are towns with populations highly vaccinated against other pathogens, yet only 2% of 200 individuals (4 individuals) showed seropositivity in this study. This low seropositivity corelated well with the low level of reported COVID-19 in those towns at the time and hinted that the rates observed in Accra and environs were due to the SARS-CoV-2 exposure rate, but not from cross-reactivity. Taken together, there is a low likelihood that cross-reactive antibodies played a significant role in this study.

## Conclusions and Recommendations

This study highlights a relatively high level of SARS-CoV-2 infections in the Greater Accra, Central and Eastern Regions, but not in the Upper East region. Most of these infections were unreported and likely asymptomatic.

As one of the first studies with such depth and nuance, we provide some of the first evidence of low levels of symptomatic COVID-19 infection, previously only anecdotally reported. These findings imply that there is a need for increased enforcement of COVID-19 mitigation protocols and more effective public education.

Large scale cross-sectional studies of seroprevalence across Ghana and West Africa may be a practical approach for disease tracking even as vaccines are being deployed^55,56^. The dynamism observed in demographic exposure risk over time highlights the need for continuous risk and prevalence assessments to track highly transmissible disease agents like SARS-CoV-2.

Finally, resources should be mobilised to research the molecular and immunological mechanisms underlying the apparent high tolerance to COVID-19 observed in Ghana, the West African sub-region, and Africa as a whole.

## Data Availability

All raw data related to this study is stored in a locked Cabinet at the West African Centre for Cell Biology of Infectious Pathogens. Deidentified participant data is password and encryption protected. This data can be accessed after appropriate verification and authorisation has been received from the University of Ghana Computing Services.

## Ethical Approval

Procedures in this study conform with the Ghanaian Public Health Act, 2012 (Act 851) and the Data Protection Act, 2012 (Act 843). Ethical approval was received from the Ethics Board of the College of Basic and Applied Sciences, University of Ghana (ECBAS 063/19-20), and the Ethical Review Committee of the Ghana Health Service (GHS-ERC 011/03/20).

## Consent

In line with the ethical protocol above, written informed consent was obtained for all study participants. Upon consent, each participant was assigned a unique identification number (ID). This ID was used for testing and data recording, thereby delinking participant identification with the test results and questionnaire answers. All consent forms are kept in a locked cabinet accessible to only the study PI and the WACCBIP Data Manager.

## Other WACCBIP COVID-19 Team members

Kyerewaa Boateng^1^, Jerry Quaye^1,3^, Aaron Adom Manu^1^, Eugene Boateng^1,3^, Daniel Lorlorson Okpatah^1,3^, Louisa Baaba Obbeng^1^, Oforiwaa Ofori^1,3^, Peggy Afua Agypomaah Birikorang^1,3^, Simon Donkoh^1^, Sylvester Languon^1,3^, Stephen Kotei Kotey^1,3^, Alexandra Lindsey Zune Djomkam^1,3^, Adelaide Fierti^1,3^, Claudia Adzo Anyigba^1,3^, Nancy Nyakoe^1,3^, Felix Ansah^1,3^, Darius Quansah^1,3,4^, Sylvia Tawiah-Eshun^1^, Barikisu Anna Ibrahim^1^, Elizabeth Fosua^1^, Raymond Adjei^1^

## Author contributions

GAA and FM conceived the study. GAA and PKQ designed the study, wrote the SOP for the fieldwork and supervised all aspects. YB, JKM, PCO, PKQ, WKA, AJU and some members of WCT performed validation of the kit. PKQ, JKM, PCO, DO-M, AJU, WCT, FK, EA, IA, LP, RA, CK, NAB, YB, JA, PA and GAA participated in seroprevalence screening activities. KB planned the site visits and partook in screening activities. PKQ, FD and PCO analysed the data. PKQ, JKM and GAA wrote the manuscript with additional editing by DO-M,YB, OQ and NN. All authors read and approved the final version of the manuscript before submission.

## Acknowledgements

We are sincerely grateful to all study participants for their contributions. We also thank the leadership of the various malls, markets, medical and research centres who allowed screening on their premises. We are grateful to the WACCBIP staff, especially the public engagement team, for their assistance in planning the screenings at the study sites.

## Supplemental Files

**Figure S1.**
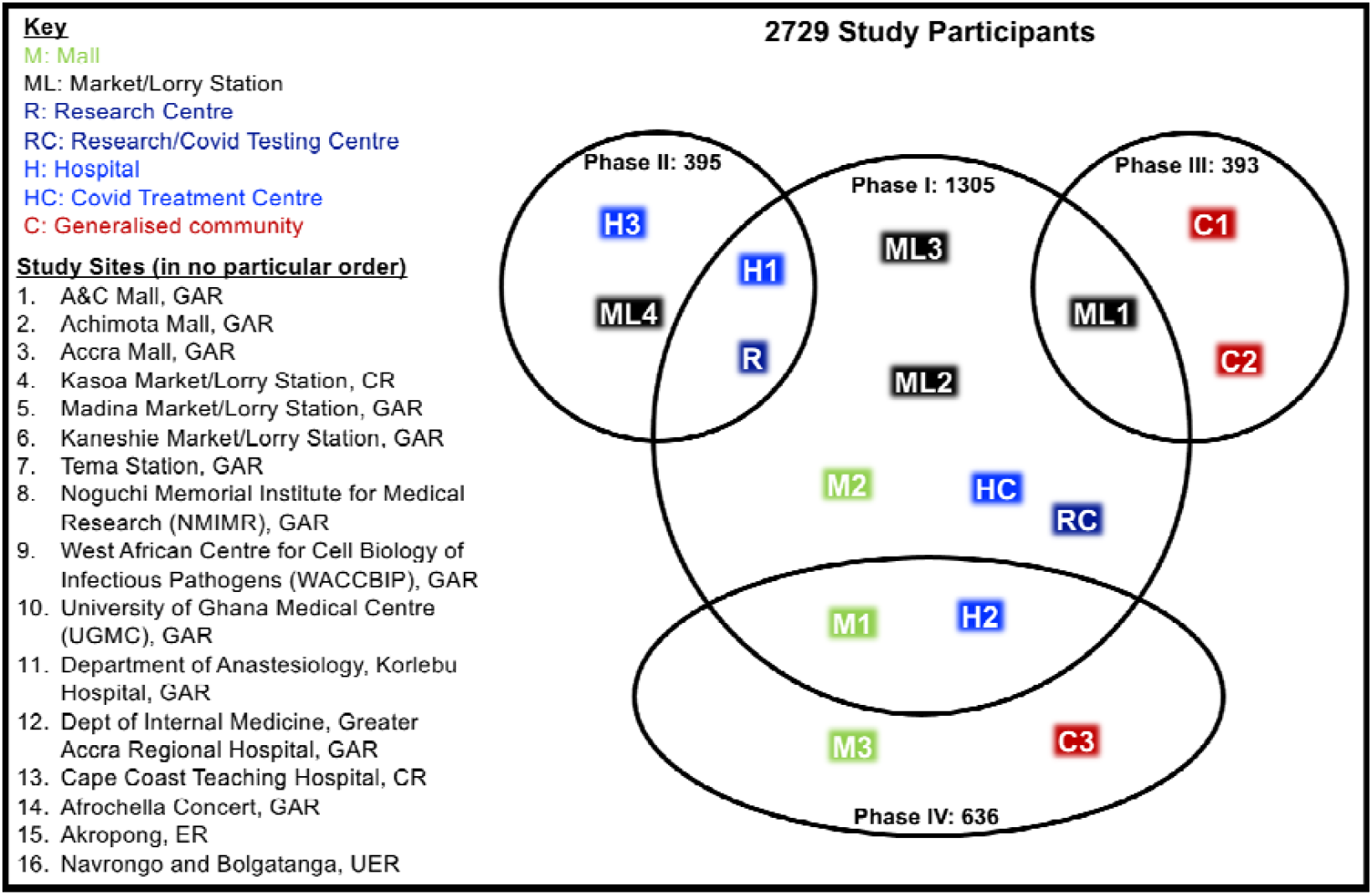
Venn Diagram showing distribution of participants and sites across the different Phases of the study. GAR refers to the Greater Accra Region, CR refers to the Central Region and UER refers to the Upper East Region.

**Figure S2:**
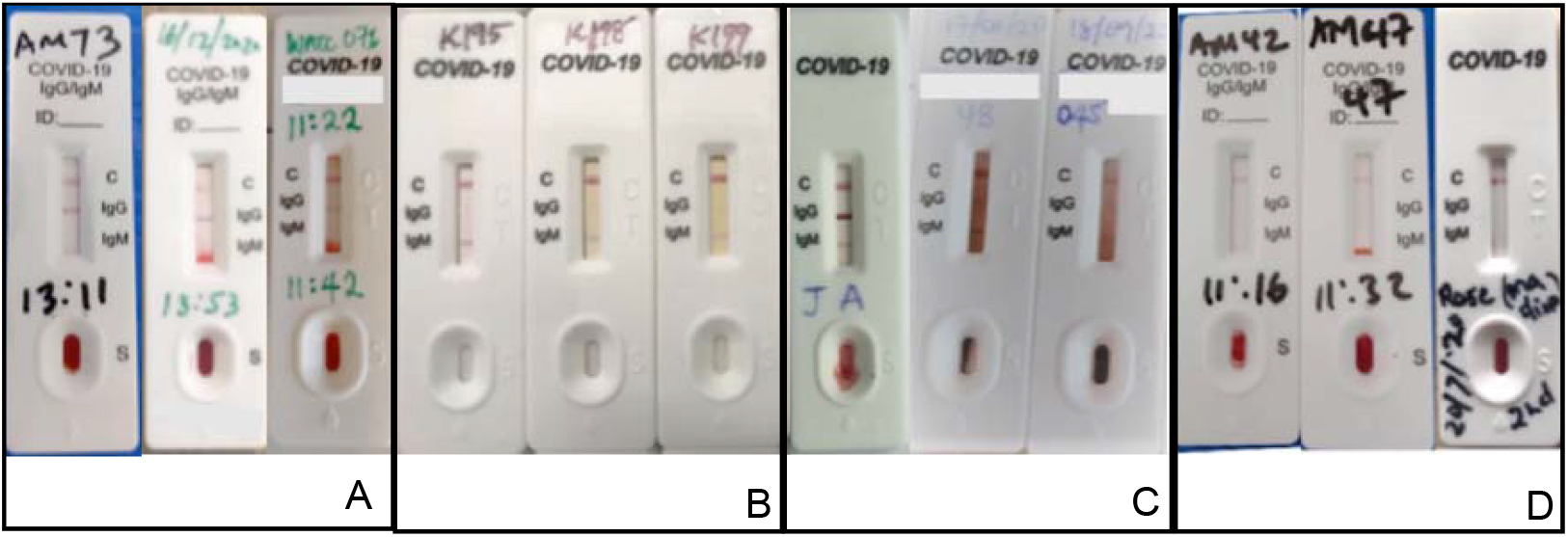
Representative pictures of study cassettes showing (A) Positive IgG, (B) Positive IgM*, (C) Combined positive IgM/IgG and (D) antibody negative test results. *Pictures of IgM are from patients, not from field due to low incidence of IgM only observed in the field and faintness of igM bands in the field

**Figure S3:**
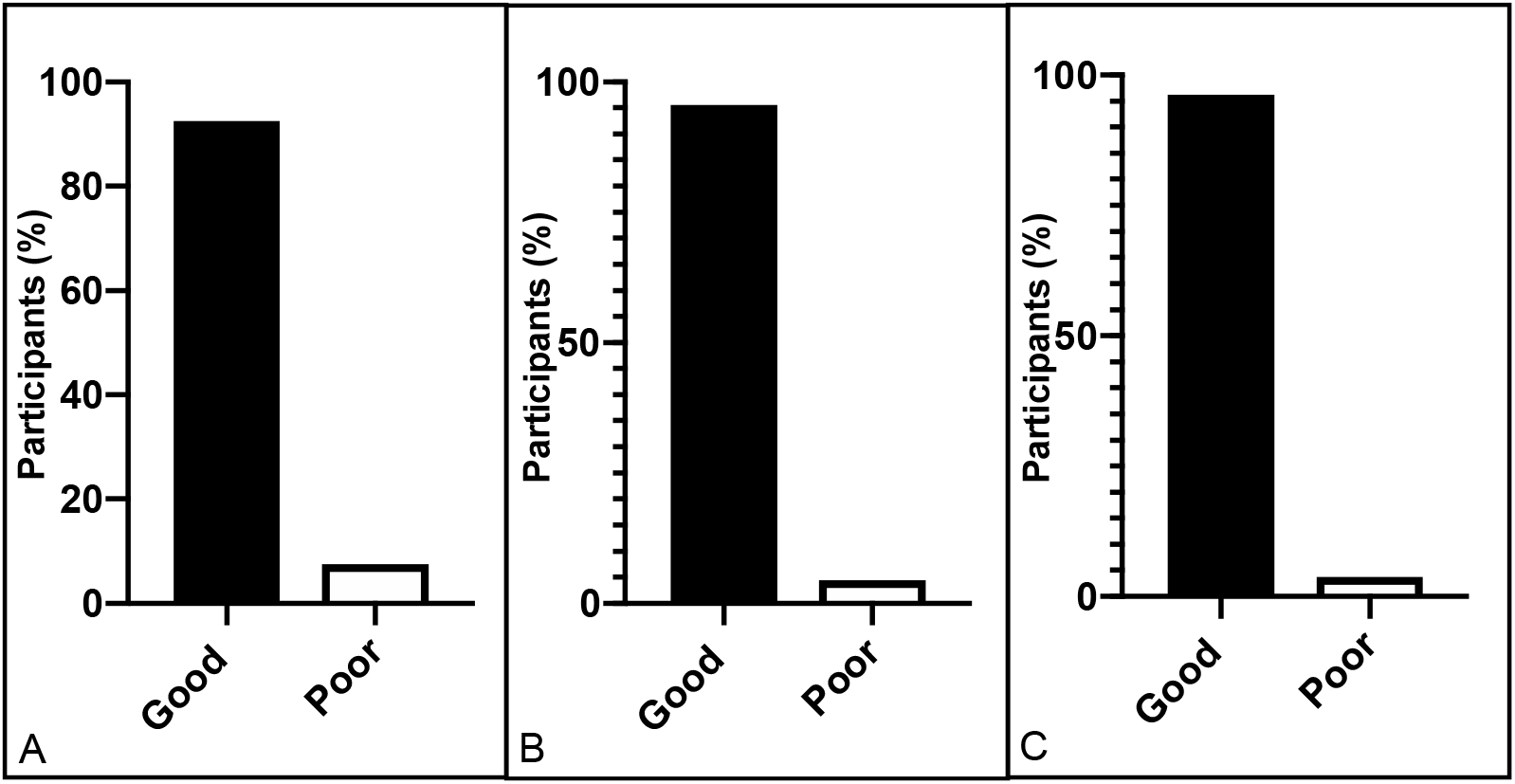
Summary of Phase I participants’ knowledge of (A) COVID-19 symptoms, (B) mode of transmission and (C) prevention.

**Figure S4:**
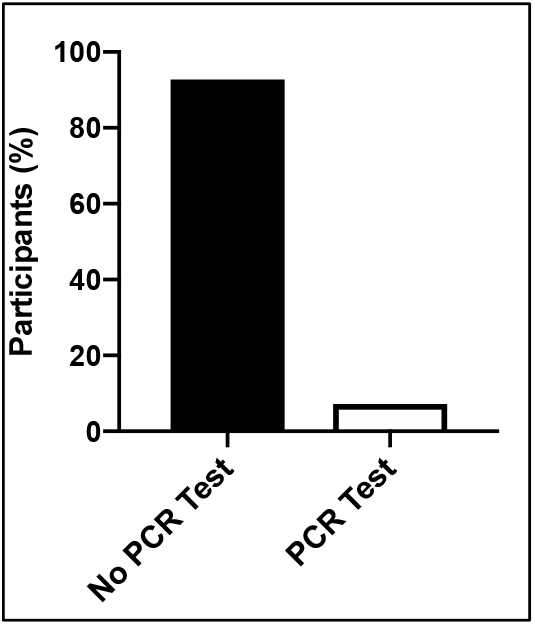
Participants who had previously taken a PCR test that detects SARS-CoV-2.

**Figure S4:**
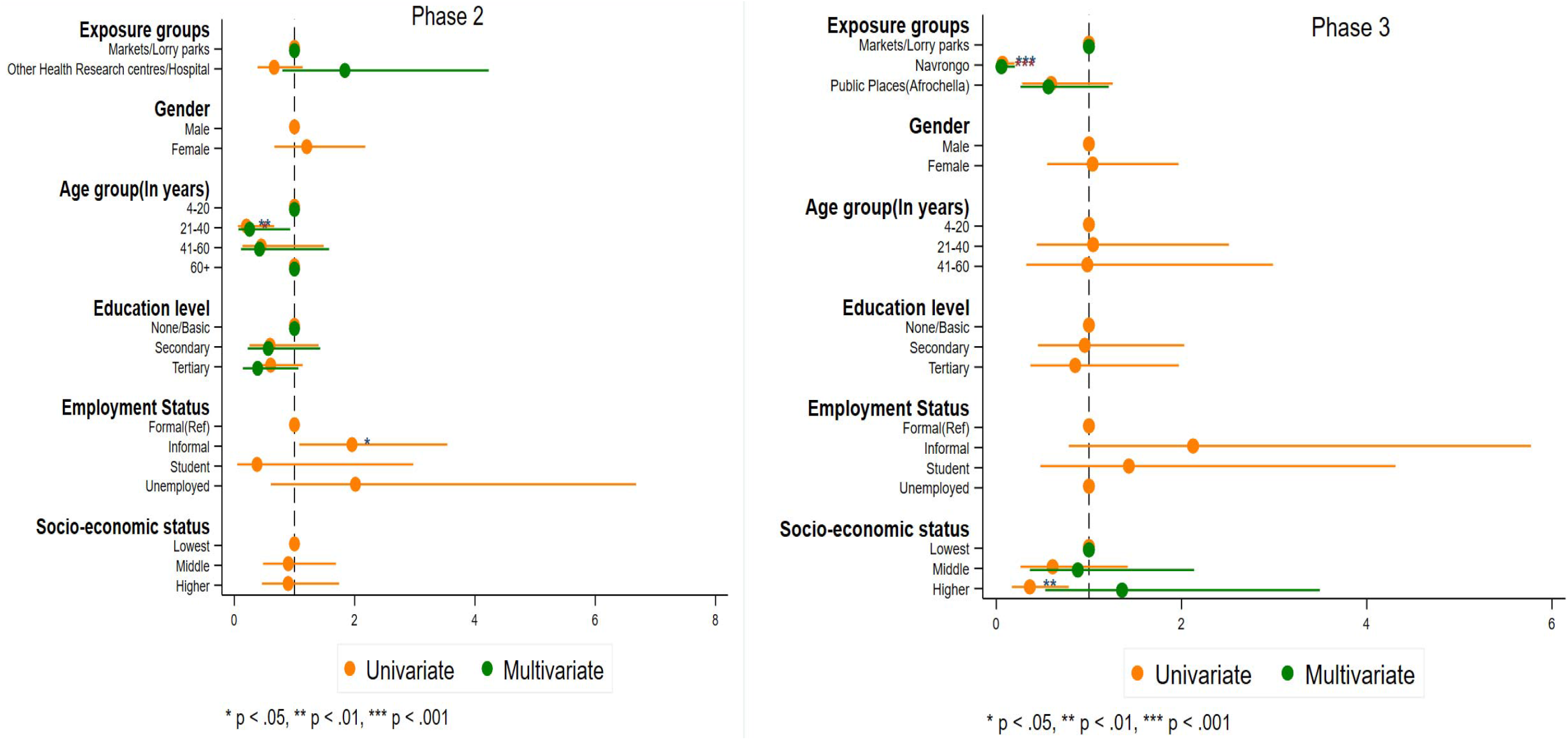
Regressional analysis of Phase II and III exposure rates.

**Figure S5:**
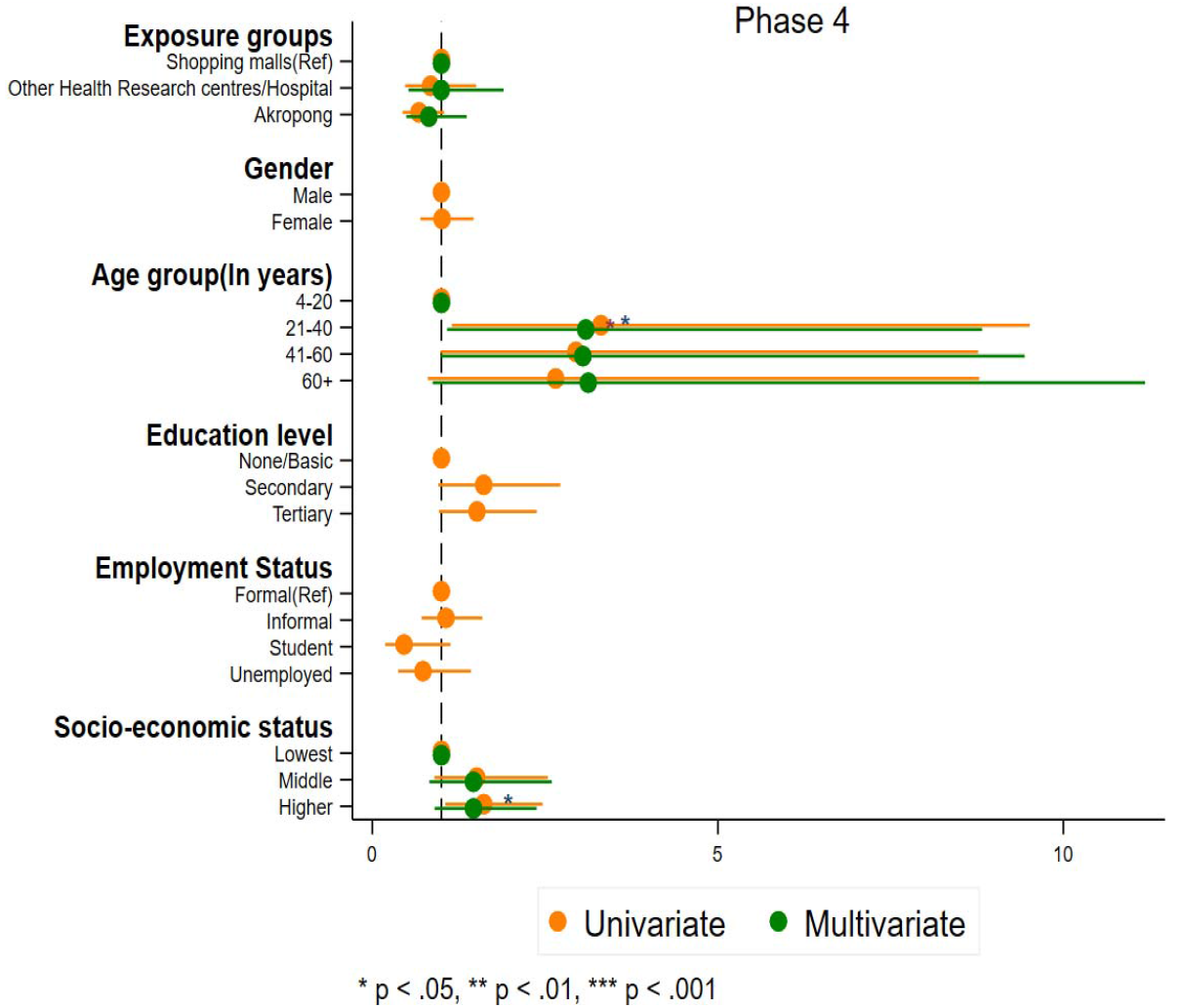
Regressional analysis of results of Phase IV exposure rates.

